# The efficacy of transcranial magnetic stimulation (TMS) for negative symptoms in schizophrenia: A systematic review and meta-analysis

**DOI:** 10.1101/2021.11.05.21265787

**Authors:** Rasmus Lorentzen, Tuan D. Nguyen, Alexander McGirr, Fredrik Hieronymus, Søren D. Østergaard

**Affiliations:** Department of Affective Disorders, Aarhus University Hospital – Psychiatry, Aarhus, Denmark; Department of Clinical Medicine, Aarhus University, Aarhus, Denmark; Hotchkiss Brain Institute, University of Calgary, Calgary, Canada; Department of Psychiatry, Cumming School of Medicine, University of Calgary, Calgary, Canada; Mathison Centre for Mental Health Research and Education, University of Calgary, Calgary, Canada; Department of Pharmacology, Sahlgrenska Academy, University of Gothenburg, Gothenburg, Sweden

## Abstract

Several trials have shown preliminary evidence for the efficacy of Transcranial Magnetic Stimulation (TMS) as a treatment for negative symptoms in schizophrenia. Here, we synthesize this literature in a systematic review and quantitative meta-analysis of double-blind randomized controlled trials of TMS in patients with schizophrenia. Specifically, MEDLINE, EMBASE, Web of Science, and PsycINFO were searched for sham-controlled, randomized trials of TMS among patients with schizophrenia. The standardized mean difference (SMD, Cohen’s *d*) with 95% confidence intervals (CI) was calculated for each study (TMS vs. sham) and pooled across studies using an inverse variance random effects model. We identified 56 studies with a total of 2550 participants that were included in the meta-analysis. The pooled analysis showed statistically significant superiority of TMS (SMD=0.37, 95%CI: 0.23; 0.52, p-value <0.00001), corresponding to a number needed to treat of 5. Furthermore, stratified analyses suggested that TMS targeting the left dorsolateral prefrontal cortex, using a stimulation frequency >1 Hz, and a stimulation intensity at or above the motor threshold, was most efficacious. There was, however, substantial heterogeneity and high risk of bias among the included studies. In conclusion, TMS appears to be an efficacious treatment option for patients with schizophrenia suffering from negative symptoms, but the optimal TMS parameters have yet to be resolved.

## Introduction

Pharmacological treatment is the cornerstone of care in schizophrenia and other psychotic disorders [1]. Though positive symptoms (e.g., delusions and hallucinations) respond relatively well to pharmacological treatment, negative symptoms often do not respond to the same degree [2, 3]. Indeed, patients with predominantly negative symptoms are more resistant to treatment than patients with primarily positive symptoms, and negative symptoms are strongly associated with low daily functioning and poor long-term prognosis [4-6]. Therefore, identification and development of efficacious treatments of negative symptoms is a priority [3, 7].

Transcranial magnetic stimulation (TMS) is a non-invasive treatment modality, which has been pursued in schizophrenia to treat both positive and negative symptoms, although with different methodologies and targets. Since the most recent reviews and meta-analyses studies on this topic [8-10], several new trials have emerged [11-19], with some using novel stimulation parameters as well as neuronavigation to better target key neurological structures, thereby possibly improving treatment outcomes. Here, we synthesize this literature in an up-to-date systematic review and quantitative meta-analysis of double-blind randomized controlled trials reporting on the efficacy of rTMS in the treatment of negative symptoms among patients with schizophrenia.

## Methods

### Protocol and Registration

The study protocol was registered at the International Prospective Register of Systematic Reviews (PROSPERO, ID: CRD42021238828) [20] and carried out in accordance with the Preferred Reporting Items for Systematic Reviews and Meta-Analyses (PRISMA) guidelines [21].

### Information Sources and Screening

MEDLINE (PubMed), PsycINFO, Web of Science and EMBASE were searched for relevant studies. Earlier reviews on the subject, clinicaltrials.gov, as well as citations of included studies were reviewed in order to find further eligible studies. The search was carried out on May 1^st^ 2021 using the following search string in MEDLINE: *(“schizophreni*” OR “schizoaffective disorder” OR “schizophreniform disorder” OR “schizophrenia”[MeSH Terms] OR “negative symptom*” OR “CHR” OR “Clinical High Risk” OR “Ultra High Risk” OR “UHR” OR “Psychotic Disorders”[MeSH Terms] OR “Psychotic Disorder*”) AND (“transcranial magnetic stimulation” OR “TMS” OR “rTMS” OR “theta burst” OR “iTBS” OR “cTBS” OR Transcranial Magnetic Stimulation*”[MeSH Terms])*. The analogue search strings used for the other databases are available in the Supplementary Material.

Titles and abstracts of studies identified via the search strategy described above were screened independently by two authors (RL and TDN) assisted by Covidence [22]. Full text versions of the studies deemed relevant after initial screening were subsequently assessed for eligibility.

### Eligibility Criteria

The following inclusion criteria were employed:

- Randomized, sham-controlled trials of transcranial magnetic stimulation (e.g. rTMS or theta burst stimulation)
- Participants with a primary diagnosis of schizophrenia, schizoaffective disorder, schizophreniform disorder or another psychotic disorder according to the DSM-IV, DSM-5, or ICD-10.
- Adult participants (≥18 years)
- Outcome measured using an established psychometric scale for negative symptoms in schizophrenia (e.g., the negative subscale of the Positive and Negative Syndromes Scale (PANSS-N) [23] or the Scale for Assessment of Negative Symptoms (SANS) [24].

The following exclusion criterion was employed:

- Co-initiation of other treatments, e.g. pharmacological treatment, as the results of such studies could be affected by an interaction effect between TMS and the co-initiated treatment.

### Data extraction

The following items were extracted from each included study: Author name, publication year, country, study type (cross-over or parallel), analysis-type (per protocol or intention-to-treat (ITT)), number of participants, drop-out rates, mean age of participants, sex distribution of participants, diagnosis, whether samples were selected for predominantly negative symptoms, frequency and intensity of TMS including the total number of stimuli and number of treatments, TMS target, nature of the sham intervention, outcome measure (rating scale), post treatment scores, follow-up scores and post treatment depression scores, if available.

If these data were not reported, the authors were contacted by e-mail with a request to provide the data. If authors did not reply, data from graphs (if available) were extracted using the GetData Graph Digitizer [25]. Previous meta-analyses were screened for post-treatment outcome data required to compute effect sizes. Studies where data was not available upon request, via graphs or through previous meta-analyses, were excluded from the analyses.

### Evaluation of risk of bias

Using the Cochrane Risk of Bias Tool 2.0 [26], the included studies were evaluated according to five domains of bias (articles in non-English languages were not evaluated): A) Randomization process, B) Deviations from intended interventions, C) Missing outcome data, D) Measurement of the outcome, and E) Selection of the reported result. The highest risk score assigned in one of these domains defined the overall risk of bias score for the study. Furthermore, potential publication bias was explored using a funnel plot.

### Statistical Analysis

The standardized mean difference between TMS and sham (SMD, Cohen’s *d*) with 95% confidence intervals (CI) was calculated for each study. SMDs were calculated based on endpoint scores or change scores for each group, with endpoint scores being preferred. If multiple outcome measures were used, PANSS-N was preferred. If a study did not provide standard deviations (SD) or data that could be used to calculate SD (e.g. standard error), the mean standard deviation across all studies of the same outcome measure was used. For cross-over studies, data was extracted after the first treatment phase (before cross-over) to exclude possible carry-over effects of treatment and thus regarded as a parallel design study.

SMDs were pooled using the inverse variance random effects model in Review Manager 5.3 [27]. This model takes into account both in-study and between-study variability. For the primary analysis, number needed to treat (NNT) was estimated using the method proposed by Kraemer and Kupfer [28]. Heterogeneity was assessed using the *I* ^2^-test with *I* ^2^-values ≥50% suggesting considerable heterogeneity. For subgroup analyses with heterogeneity <50%, the fixed effect model was used for analysis. For multi-arm studies, data from different active TMS treatment arms were pooled in the calculation of overall efficacy as to not duplicate data from the sham group, using the formulas provided in table 6.5a in the Cochrane Handbook [29].

Following the main analysis, separate effect size analyses were carried out i) after stratifying by type of TMS, ii) after stratifying by stimulation frequencies, iii) after stratifying by stimulation intensity, iv) for patients with predominantly negative symptoms, v) after excluding studies with data extracted from graphs, vi) after excluding studies with high risk of bias. vii) after stratifying by age, viii) after excluding studies reporting change-from-baseline scores, ix) focusing on long term effect using data from at least four weeks after the last treatment (the last follow up in each study was used), x) focusing on the effect size of TMS for depressive symptoms (all depression measures allowed with a preference for the Calgary Depression Scale in Schizophrenia [30]), and finally xi) after excluding studies with data extracted from other reviews.

## Results

### Study Selection

The search yielded 3287 articles of which 1573 were duplicates, resulting in 1714 studies that underwent title- and abstract screening (Figure 1). Following this screening, 1565 were excluded. This left 149 articles to be assessed in full text, of which 80 studies did not meet the eligibility criteria. Of the 69 eligible studies, 35 reported insufficient data and thus the authors were contacted by e-mail requesting additional data. The means and standard deviations of PANSS-N was the most common missing piece of information (i.e., from studies where only the total PANSS scores were reported). From these 35 studies, three author groups provided data [13, 18, 31], and data were extracted from graphs in an additional eight studies [19, 32-38]. Hence, 24 articles were excluded due to non-available data [39-62]. In total, the search yielded 45 includable studies, with 51 comparisons as a result of studies including multiple interventions [11-19, 31-38, 63-90]. No additional studies were found in citations or in the database of clinicaltrials.gov. Summary data was available from 14 studies reviewed by Wang and colleagues [10] from non-English reports, [87-89, 91-101], of which 3 was studies found through the database search, leaving a total of 56 studies and 62 comparisons [11-19, 31-38, 62-101].

**Figure 1:**
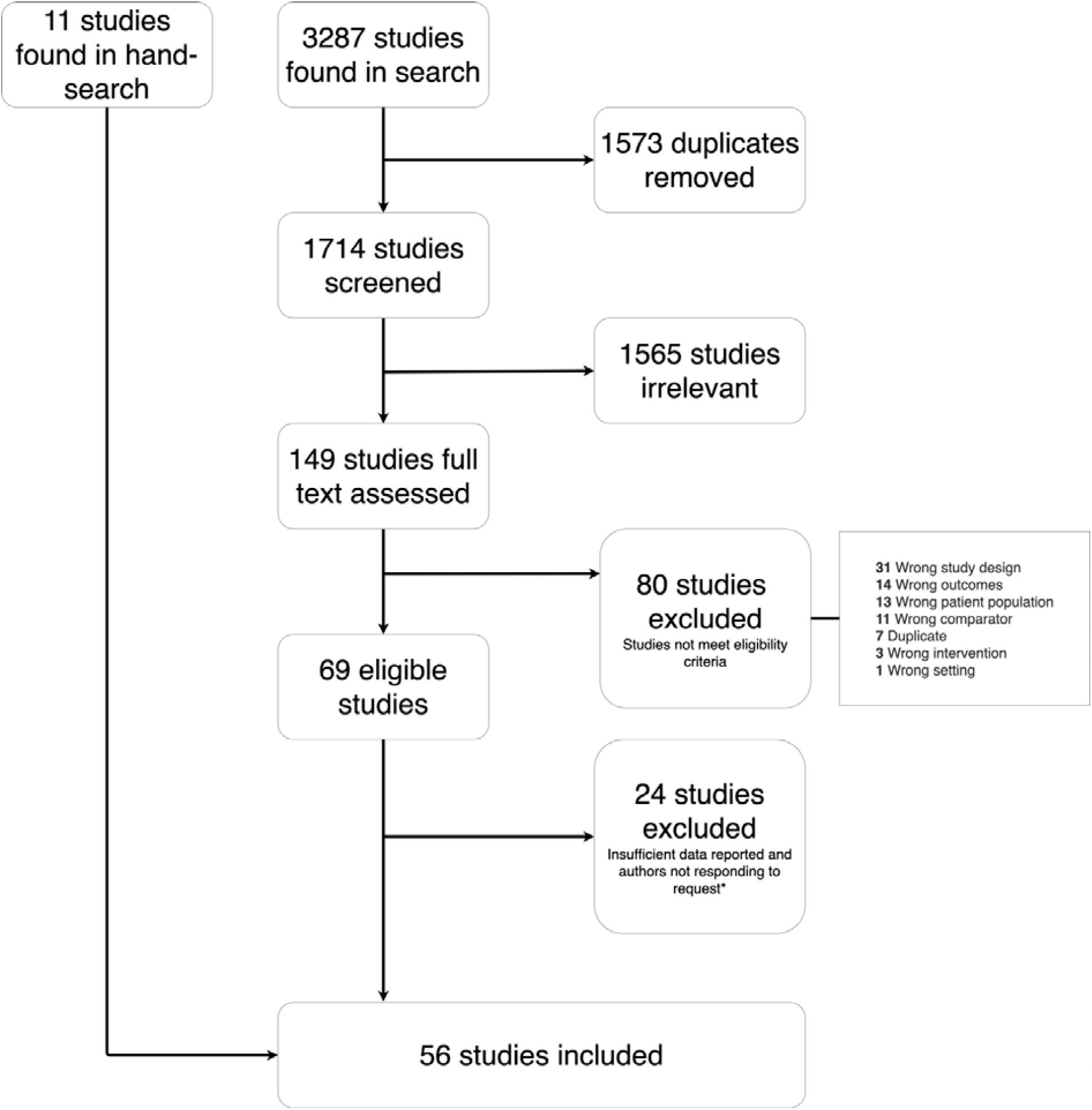
PRISMA flowchart *Authors were contacted by e-mail. If data was not provided and data could not be taken from graphs, the study was excluded.

### Study characteristics

The 56 studies included 2550 participants, of whom 1440 received active treatment and 1110 sham treatment (Table 1). In the 54 studies (*n*=2442) that reported the diagnoses of the participants, 98.8% had schizophrenia and 1.2 % had schizoaffective disorder. The studies were conducted in 15 different countries, of which China was the most common (*n*=24). Almost all included studies reported the outcome using PANSS-N or SANS with only one study using the Brief Psychiatric Rating Scale – Negative/Disorganized factor (BPRS-N/D) [65].

**Table 1:**
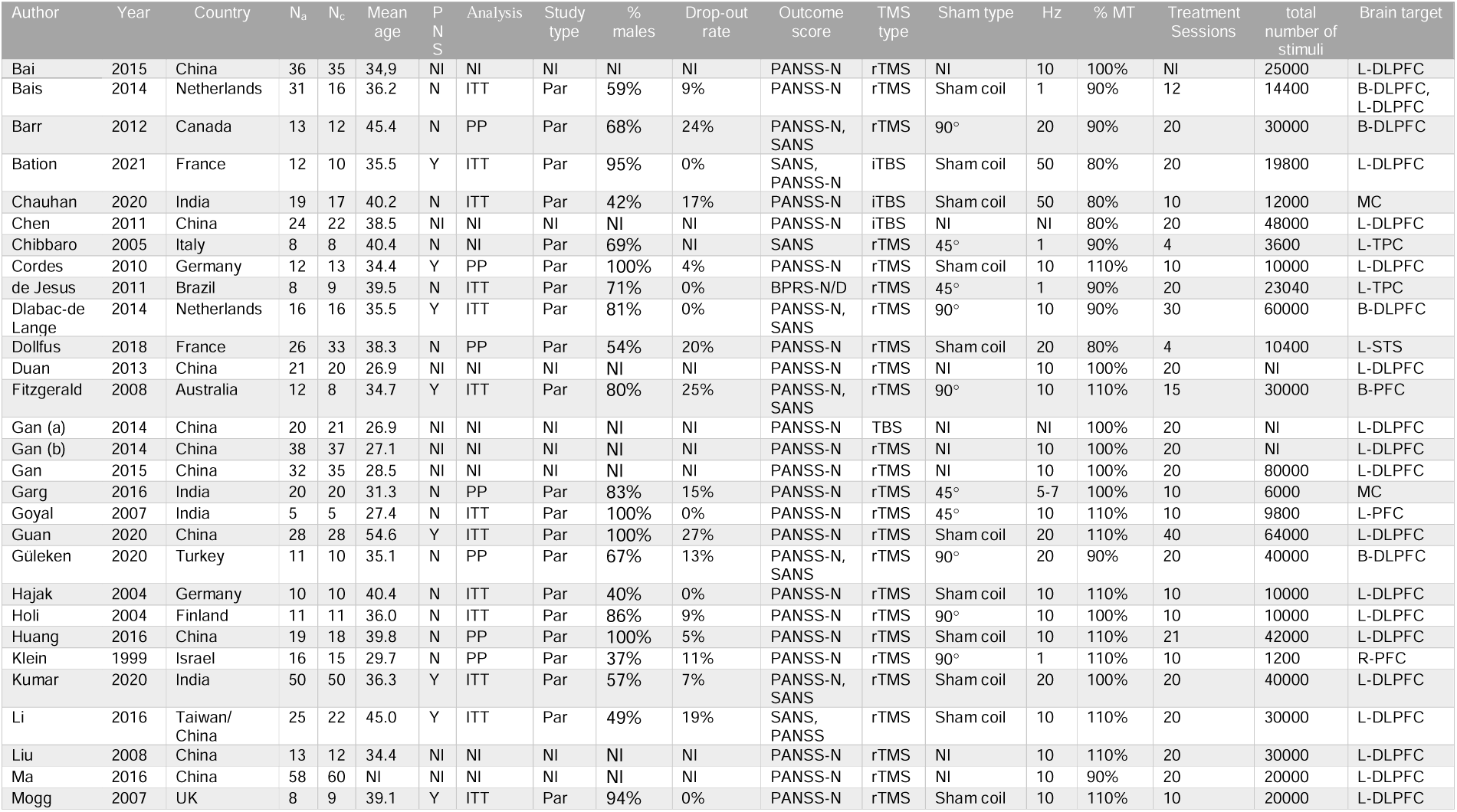
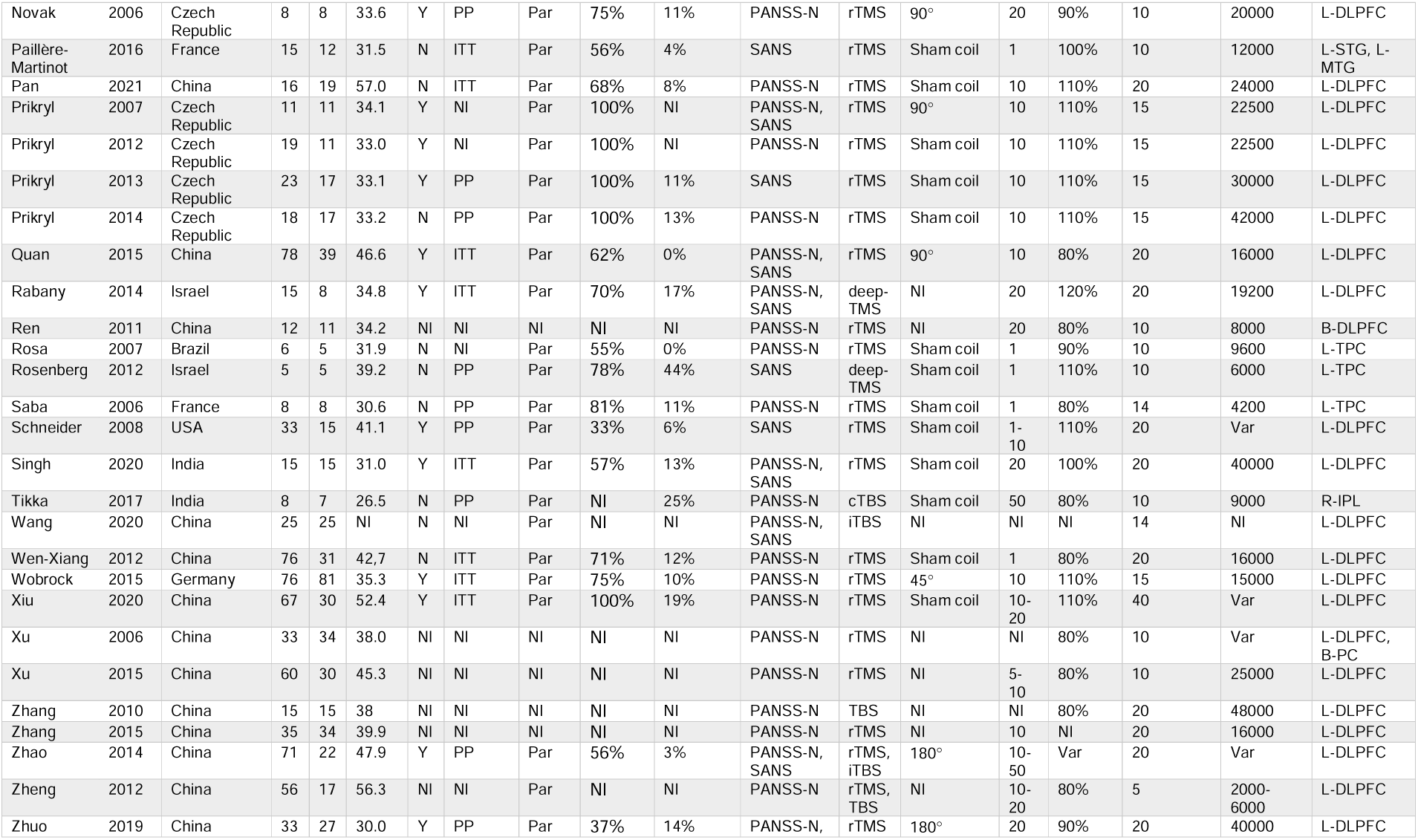
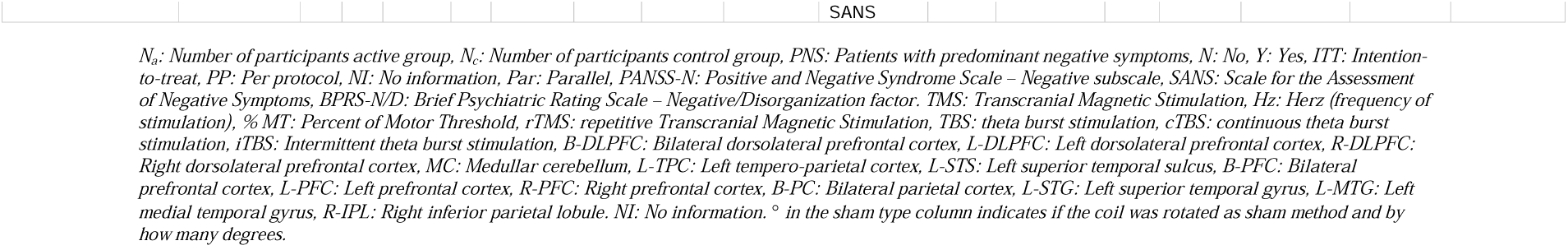
Characteristics of included studies

Several different active TMS modalities were used in the included trials, with some testing more than one active modality against sham treatment: rTMS (47 studies, 10 used ≤1 Hz, 38 used >1 Hz, 41 used unilateral treatment, and eight bilateral or midline treatment), TBS (9 studies, 5 used iTBS, 1 used cTBS, and 3 used unspecified TBS), and deep-TMS (2 studies) (see Supplementary Table 2, treatment characteristics). The mean total number of stimuli per trial was 25,684 varying from 1200-80,000 stimuli. The majority of the studies (*n*=39) had the left dorsolateral prefrontal cortex (L-DLPFC) as the primary stimulation target.

### Risk of Bias of Individual Studies

Eight studies were regarded as having low risk of bias, 10 studies with “some concerns”, and 23 studies with high risk of bias (Supplementary Table 3). The most common reason (n=23) for “some concerns” was insufficient reporting whether the randomization sequence was concealed adequately (domain A). Improper analysis (e.g. “per protocol” analysis, domain B) and missing outcome data (domain C) were the most common reasons (n=12 and n=21, respectively, with n=11 having both) for a study being regarded as having high risk of bias.

### Results of Individual Studies

Standardized mean differences for the included studies are shown in Figure 2. Seventeen studies showed a statistically significant superior effect of TMS compared to sham treatment [13, 14, 34, 75, 76, 78, 79, 85-87, 91, 92, 96-99, 101] and one study found a statistically significant superior effect of sham treatment [32]. The remaining studies did not show a statistically significant difference between the treatment groups. There was considerable heterogeneity between the included studies (*I*^2^ = 65%).

**Figure 2:**
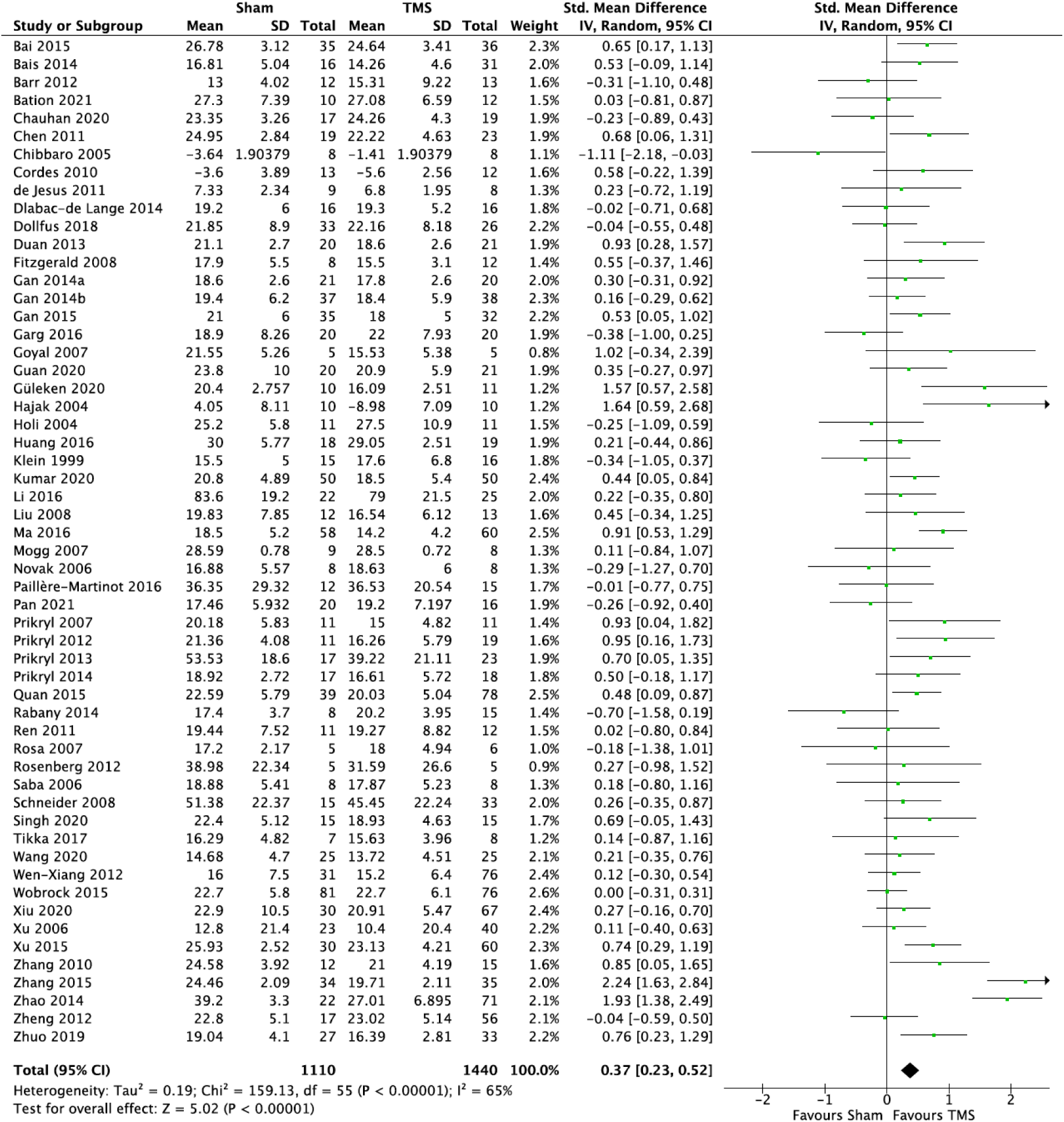
Forest plot of standardized mean differences (effect size)

### Synthesis of Results

As evident from Figure 2, the overall SMD was 0.37 (95%CI: 0.23; 0.52, p < 0.00001) in favor of TMS, corresponding to an NNT of 5. The results of the secondary analyses are available in Table 2. Using follow-up data from at least four weeks after end of treatment yielded an SMD of 0.25 (95%CI: 0.09; 0.40). Following exclusion of i) studies with data taken from graphs (SMD=0.40, 95%CI: 0.24; 0.56), ii) studies with a high risk of bias (SMD=0.35, 95%CI: 0.08; 0.62), or iii) studies using change-from-baseline scores (SMD=0.38, 95%CI: 0.24; 0.53) did not impact the effect estimate substantially. Stimulation of L-DLPFC seemed to have a larger effect than other sites (SMD=0.51, 95%CI: 0.33; 0.69 vs. SMD=0.05, 95%CI: -0.14; 0.23), however, there was considerable methodological heterogeneity in the “other” category. The SMD of TBS (SMD=0.48, 95%CI: -0.01; 0.96) was similar to that of rTMS (SMD=0.46, 95%CI: 0.27; 0.65). The effect in participants with predominantly negative symptoms was highly significant (SMD=0.45, 95%CI: 0.20; 0.70), while no effect on depressive symptoms was observed (SMD=0.03, 95%CI: -0.12; 0.18.

**Table 2:** Results of overall effect size and subgroup analyses

|  |  | *N* | SMD (95% confidence interval) | *p* | *I*<sup>2</sup> |
| --- | --- | --- | --- | --- | --- |
| Overall standardized mean difference (SMD) |  | 2550 | 0.37 (0.23, 0.52) | **<0.00001** | 65% |
| Site | L-DLPFC | 2072 | 0.51 (0.33, 0.69) | **<0.00001** | 71% |
|  | Other* | 490 | 0.05 (-0.14, 0.23) | 0.62 | 24% |
| Stimulation Frequency (rTMS only) | 1 Hz* | 304 | 0.05 (-0.18, 0.29) | 0.66 | 5% |
|  | > 1 Hz | 1853 | 0.52 (0.42, 0.62) | **<0.00001** | 78% |
| Stimulation intensity | <100% of MT | 1026 | 0.26 (0.06, 0.46) | **0.01** | 53% |
|  | ≥100% of MT | 1427 | 0.42 (0.23, 0.61) | **<0.0001** | 63% |
| Age** | ≤ median* | 1026 | 0.33 (0.20, 0.45) | **<0.00001** | 46% |
|  | > median | 1352 | 0.40 (0.14, 0.66) | **0.002** | 78% |
| Type | rTMS | 2218 | 0.47 (0.28, 0.66) | **<0.00001** | 76% |
|  | TBS | 352 | 0.48 (-0.01, 0.96) | 0.06 | 78% |
|  | Deep-TMS* | 33 | -0.34 (-1.07, 0.38) | 0.36 | 47% |
| Excluding data from graphs |  | 2365 | 0.40 (0.24, 0.56) | **<0.00001** | 69% |
| Excluding high risk of Bias studies*** |  | 813 | 0.35 (0.08, 0.62) | **0.01** | 69% |
| Excluding change-from-baseline studies |  | 2548 | 0.38 (0.24, 0.53) | **<0.00001** | 66% |
| Participants with PNS |  | 1034 | 0.45 (0.20, 0.70) | **0.0005** | 69% |
| Follow up (≥ 4 weeks after end of treatment*) |  | 695 | 0.25 (0.09, 0.40) | **0.002** | 42% |
| Depressive symptoms* |  | 743 | 0.03 (-0.12, 0.18) | 0.72 | 30% |
| Excluding data taken from earlier reviews |  | 1784 | 0.29 (0.12, 0.45) | **0.0006** | 60% |

*L-DLPFC: Left dorsolateral prefrontal cortex, rTMS: repetitive transcranial magnetic stimulation, MT: motor threshold, TBS: Theta burst stimulation, deep-TMS: deep transcranial magnetic stimulation, PNS: Predominant negative symptoms. \*calculated using fixed effect analysis due to heterogeneity <50%, \*\*median = 35,5 years, \*\*\*Non-English language articles also excluded as they were not evaluated. Some studies had several active treatment groups and are included in several analyses.*

### Risk of Bias Across Studies

Based on the funnel plot (Figure 3), two outlying studies suggest some degree of asymmetry, indicating a small to moderate risk of publication bias.

**Figure 3:**
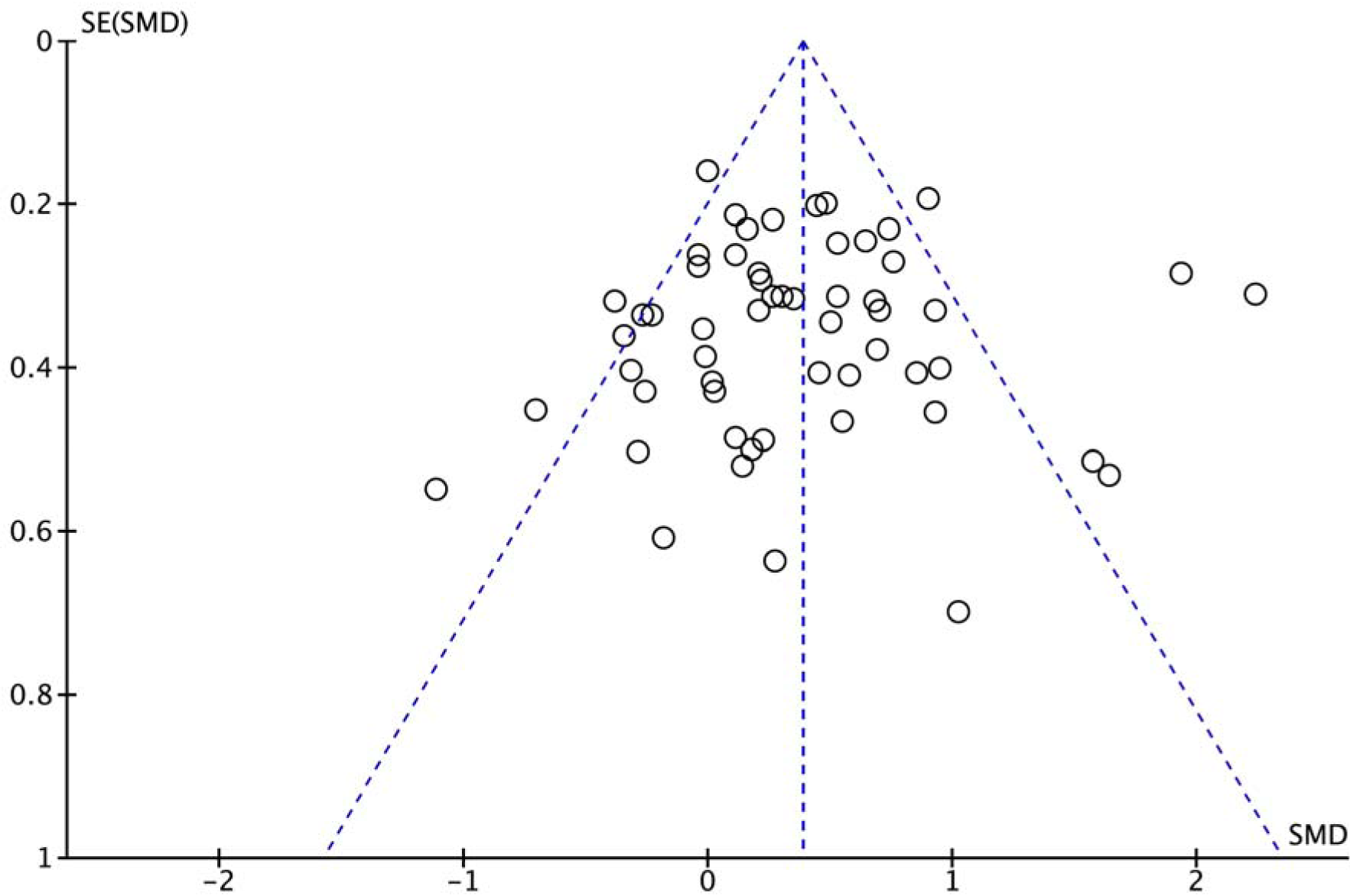
Funnel plot SE: Standard error, SMD: Standardized mean difference

## Discussion

Based on meta-analysis of 56 studies with a total of 2550 participants mainly with schizophrenia, we found a superior effect of active TMS on negative symptoms compared to sham treatment. The SMD was 0.37 (95%CI: 0.23; 0.52) in favor of active TMS, translating to an NNT of 5. The superiority of active TMS remained statistically significant following a) exclusion of data extracted from graphs, b) exclusion of studies deemed to be at high risk of bias, and c) exclusion of studies reporting change-from-baseline scores. Further subgroup analyses suggested that using >1 Hz stimulation (SMD=0.52 vs. SMD=0.05), targeting the L-DLPFC (SMD=0.51 vs. SMD=0.05), and ≥100% motor threshold (SMD=0.42 vs. SMD=0.26) may be more effective. However, there was considerable heterogeneity across the included studies and these results should be considered tentative. In contrast to a prior meta-analysis [8], we found no support for the suggestion that TMS should have particularly beneficial effect upon negative symptoms for younger individuals (SMD=0.33 vs. SMD=0.40).

While there are prior meta-analysis on this topic, the present work covers substantially more participants (77% more than Wang et al. [10]) and studies (80% more studies than Osoegawa et al. [9]). Both of these prior meta-analyses also pointed to a beneficial effect of TMS on the negative symptoms of schizophrenia. This up-to-date meta-analysis therefore consolidates the sentiment that there may be a place for TMS in the treatment of negative symptoms in schizophrenia, which have a highly adverse effect on daily function and are often resistant to other types of treatment [5].

Several different brain areas were targeted by the studies included in this synthesis, in which subgroup analyses suggested that stimulation of the L-DLPFC had superior effect compared to other targets (SMD=0.51 vs. SMD=0.05). These results align with earlier studies that have found an inverse correlation between frontal lobe size and glucose metabolism, and negative symptom severity [102, 103]. Moreover, there are increasing data suggesting that the DLPFC has a privileged relationship with other structures implicated in negative symptoms, including the midline cerebellum (MC) [104]. The circuitry of the DLPFC in relation to negative symptoms will likely be an important area to study when developing stimulation targets in personalized medicine.

There are limitations to this study, which should be acknowledged by the readers. First, as there are phenomenological overlaps between negative and depressive symptoms and since depression responds well to TMS [105, 106], the relief of depressive symptoms during treatment could potentially confound the estimation of the effect on negative symptoms. However, our analysis of data from studies measuring depressive symptoms in the context of schizophrenia found no statistically significant efficacy regarding depressive symptoms, suggesting that the effect on negative symptoms is not confounded in this respect. Second, 56% of the evaluated studies were regarded as “high risk of bias” studies, which is a substantially larger proportion compared to the 13% reported in the review by Wang et al [10]. This difference is predominantly a consequence of classification as we used the Cochrane Risk of Bias Tool 2.0, while Wang et al. used the Cochrane Risk of Bias Tool 1.0. The most common reason for studies being considered as “high risk” in the context of the present review was missing data. We employed a relatively conservative 10% cut-off for missing data, but there is no agreed upon threshold [107] and the proportion of studies classified as “high risk of bias” can thus vary considerably between reviews. Third, we used a broad search strategy, but relevant studies may have been missed nevertheless. However, assuming that such potential misses occur at random, it should not have affected the reported efficacy estimates. Fourth, the inclusion of data drawn from reviews is suboptimal. However, the analysis excluding this data yielded results equivalent to those from the primary analysis. Fifth, while we conducted several subgroup analyses, formal (effect) moderator analysis was not conducted. Sixth, there was significant heterogeneity in outcome across the included studies, with *I* ^2^-assessments at 50% or above in all but six cases (65% in the primary analysis; Figure 2). While this is likely partly due to the considerable heterogeneity of the TMS treatment provided across the included studies, other sources of heterogeneity, such as differences in sham conditions, patient populations, outcome measures, or random chance, are also likely contributors. Relatedly, in the review by He et al., a univariate meta-regression of stimulation frequency, total simulation, motor threshold, stimulation site, study design, and type of coil was conducted. None of these factors were shown to be the main source of heterogeneity.

In conclusion, this systematic review and quantitative synthesis of sham-controlled studies suggests that TMS is efficacious in the treatment of negative symptoms of schizophrenia. Although it appears that targeting the L-DLPFC, using a stimulation frequency >1 Hz, and stimulation intensity at or above the motor threshold are the most efficacious settings, the optimal treatment parameters are yet to be resolved.

## Data Availability

All data produced in the present work are contained in the manuscript and supplementary files.

