## Supplementary Material for "The efficacy of transcranial magnetic stimulation (TMS) for negative symptoms in schizophrenia: A systematic review and meta-analysis"

### **Appendix**

Supplementary Table 1: Follow up data

| Author | Year | Follow up time | Follow up score (SD)_active_ | | Follow up score (SD)_control_ | |
| --- | --- | --- | --- | --- | --- | --- |
| Bation | 2021 | 6 months | 22.83 | (6.043) | 27.3 | (10.38) |
| Chibbaro | 2005 | 8 weeks | -8.28 | (2.72) | 0.45 | (0.19) |
| de Jesus | 2011 | 4 weeks | 9.5 | (3.81) | 12.56 | (4.79) |
| Dlabac-de Lange | 2014 | 3 months | 18.1 | (4.6) | 18.2 | (6) |
| Dollfus | 2018 | 4 weeks | 22.77 | (7.81) | 22.97 | (8.39) |
| Li | 2016 | 4 months | 15.7 | (4.93) | 19.3 | (5.92) |
| Liu | 2008 | 4 weeks | 72.5 | (16.8) | 83.5 | (20.5) |
| Novak | 2006 | 6 weeks | 18 | (5.73) | 15.38 | (5.1) |
| Rabany | 2014 | 4 weeks | 18.6 | (4.93) | 15 | (5.83) |
| Rosa | 2007 | 4 weeks | 18 | (4.8) | 18.75 | (0.5) |
| Schneider | 2008 | 4 weeks | 46.59 | (22.55) | 50.93 | (22.37) |
| Wang | 2020 | 2 months | 12.08 | (3.66) | 14.52 | (4.78) |
| Wobrock | 2015 | 84 days | 19.99 | (50.82) | 20.32 | (63) |
| Xiu | 2020 | 6 months | 20.76 | (8.48) | 22.2 | (7.9) |

*SD: Standard deviation*

Supplementary Table 2: Data on depressive symptoms

| Author | Year | Depression scale used | Post treatment depression score (SD)_active_ | | Post treatment depression score (SD)_control_ | |
| --- | --- | --- | --- | --- | --- | --- |
| Barr | 2012 | CDSS | 2.23 | (1.87) | 1.58 | (2.5) |
| de Jesus | 2011 | BPRS-DF | 3.37 | (2.26) | 3.55 | (3.24) |
| Dlabac-de Lange | 2014 | MADRS | 16.3 | (7.8) | 11.8 | (6.8) |
| Fitzgerald | 2008 | CDSS | 7.2 | (5.9) | 3.5 | (3.8) |
| Garg | 2016 | CDSS | 6.3 | (3.45) | 6.5 | (3.68) |
| Goyal | 2007 | CDSS | 0 | (0) | 0.8 | (0.836) |
| Guan | 2020 | PANSS-DF | 3.9 | (2) | 4.2 | (1.6) |
| Huang | 2016 | MADRS | 14.89 | (5.52) | 12.39 | (2.57) |
| Klein | 1999 | HDRS | 8.6 | (3.5) | 6.9 | (4) |
| Kumar | 2020 | CDSS | 0.12 | (0.44) | 0.12 | (0.72) |
| Mogg | 2007 | HADS-D | 2.5 | (3.1) | 5.2 | (3.3) |
| Prikryl | 2007 | CDSS | 0 | (0) | 0.73 | (1.19) |
| Prikryl | 2013 | CDSS | 0.04 | (0.21) | 0.76 | (1.48) |
| Prikryl | 2014 | CDSS | 0.92 | (0.78) | 1.09 | (1.51) |
| Rabany | 2014 | CDSS | 5.8 | (3.23) | 5.5 | (4.78) |
| Singh | 2020 | CDSS | 1.33 | (0.97) | 1.33 | (1.23) |
| Wang | 2020 | HDRS | 3.76 | (1.45) | 5.4 | (3:33) |
| Wobrock | 2015 | CDSS | 4.4 | (3.5) | 4.6 | (4.4) |

*CDSS: Calgary Depression Scale for Schizophrenia, BPRS-DF: Brief Psychiatric Rating Scale – depressive factor, MADRS: Montgomery-Asberg Depression Rating Scale, PANSS-DF: Positive and Negative Syndrome Scale – Depressive Factor, HDRS: Hamilton Depression Rating Scale, HADS-D: Hospital Anxiety and Depression Score – Depression score.*

Supplementary Table 3: Risk of bias in individual studies

|  | | **Cochrane Risk of Bias 2.0 domains** | | | | | **Overall score** |
| --- | --- | --- | --- | --- | --- | --- | --- |
| **Author** | **Year** | **A** | **B** | **C** | **D** | **E** |  |
| Bais | 2014 | L | L | L | L | L | **L** |
| Barr | 2012 | SC | H | H | L | L | **H** |
| Bation | 2021 | L | L | L | L | L | **L** |
| Chauhan | 2020 | L | SC | L | L | L | **SC** |
| Chibbaro | 2005 | SC | SC | H | L | L | **H** |
| Cordes | 2010 | SC | H | L | L | L | **H** |
| de Jesus | 2011 | L | L | L | L | L | **L** |
| Dlabac-de Lange | 2014 | L | L | L | L | L | **L** |
| Dollfus | 2018 | SC | H | H | L | L | **H** |
| Fitzgerald | 2008 | L | SC | H | L | L | **H** |
| Garg | 2016 | SC | H | H | L | L | **H** |
| Goyal | 2007 | H | L | L | L | L | **H** |
| Guan | 2020 | L | SC | H | L | L | **H** |
| Güleken | 2020 | H | SC | H | L | L | **H** |
| Hajak | 2004 | SC | L | L | L | L | **SC** |
| Holi | 2004 | L | L | L | L | L | **L** |
| Huang | 2016 | SC | SC | L | L | L | **SC** |
| Klein | 1999 | SC | H | H | L | L | **H** |
| Kumar | 2020 | L | L | L | L | L | **L** |
| Li | 2016 | SC | SC | L | L | L | **SC** |
| Mogg | 2007 | L | L | L | L | L | **L** |
| Novak | 2006 | SC | H | H | L | L | **H** |
| Paillère-Martinot | 2016 | L | L | L | L | L | **L** |
| Pan | 2021 | SC | L | L | L | L | **SC** |
| Prikryl | 2007 | SC | L | L | L | L | **SC** |
| Prikryl | 2012 | SC | H | H | L | L | **H** |
| Prikryl | 2013 | SC | H | H | L | L | **H** |
| Prikryl | 2014 | SC | H | H | L | L | **H** |
| Quan | 2015 | SC | SC | L | L | L | **SC** |
| Rabany | 2014 | SC | L | H | L | L | **H** |
| Rosa | 2007 | SC | L | L | L | L | **SC** |
| Rosenberg | 2012 | L | H | H | L | L | **H** |
| Saba | 2006 | SC | SC | H | L | L | **H** |
| Schneider | 2008 | SC | SC | L | L | L | **SC** |
| Singh | 2020 | L | L | H | L | L | **H** |
| Tikka | 2017 | L | H | H | L | L | **H** |
| Wang | 2020 | SC | SC | H | L | L | **H** |
| Wobrock | 2015 | L | SC | H | L | L | **H** |
| Xiu | 2020 | L | SC | H | L | L | **H** |
| Zhao | 2014 | SC | SC | L | L | L | **SC** |
| Zhuo | 2019 | SC | H | H | L | L | **H** |

*A: Randomization process, B: Deviations from intended interventions, C: Missing outcome data, D: Measurement of the outcome, E: Selection of the reported result, L: Low risk, SC: Some concerns, H: High risk.*

**Search strategy**

**PubMed:**

(“schizophreni*” OR "schizoaffective disorder" OR "schizophreniform disorder" OR "schizophrenia"[MeSH Terms] OR “negative symptom*” OR “CHR” OR “Clinical High Risk” OR “Ultra High Risk” OR “UHR” OR "Psychotic Disorders"[MeSH Terms] OR "Psychotic Disorder*") AND ("transcranial magnetic stimulation" OR "TMS" OR "rTMS" OR "theta burst" OR "iTBS" OR “cTBS” OR "transcranial Magnetic Stimulation*"[MeSH Terms])

**751 hits**

**EMBASE:**

('schizophreni*':ab,kw,ti OR 'schizoaffective disorder':ab,ti,kw OR 'schizophreniform disorder':ab,kw,ti OR 'schizophrenia spectrum disorder'/exp OR 'psychotic disorder*':ab,kw,ti OR ‘negative symptom*’:ab,kw,ti OR ‘CHR’:ab,kw,ti OR ‘Clinical High Risk’:ab,kw,ti OR ‘Ultra High Risk’:ab,kw,ti OR ‘UHR’:ab,kw,ti) AND ('transcranial magnetic stimulation':ab,kw,ti OR 'tms':ab,kw,ti OR 'rtms':ab,kw,ti OR 'theta burst':ab,kw,ti OR 'itbs':ab,kw,ti OR 'cTBS':ab,kw,ti OR 'transcranial magnetic stimulation'/exp) AND ('article'/it OR 'article in press'/it OR 'review'/it)

**902 hits**

**PsycINFO:**

(schizophreni* OR "schizoaffective disorder" OR "schizophreniform disorder" OR "Psychotic Disorder*" OR “negative symptom*” OR “CHR” OR “Clinical High Risk” OR “Ultra High Risk” OR “UHR”) AND ("transcranial magnetic stimulation" OR "TMS" OR "rTMS" OR "theta burst" OR "iTBS" OR “cTBS”)

Source type: Scholarly Journals

**699 hits**

**Web of Sciences (Web of Science, Core Database Collection):**

(schizophreni* OR "schizoaffective disorder" OR "schizophreniform disorder" OR "Psychotic Disorder*" OR “negative symptom*” OR “CHR” OR “Clinical High Risk” OR “Ultra High Risk” OR “UHR”) AND ("transcranial magnetic stimulation" OR "TMS" OR "rTMS" OR "theta burst" OR "iTBS" OR “cTBS”)

Search type: “Topic”. Document type: Article, review, proceedings paper, early acces

**935 hits**

**In total 3287 hits**

After removal of **1573** duplicates: **1714 hits**
